# Evaluating the role of liquid biopsy to detect pathogenic DNA Damage Repair (DDR) gene alterations in metastatic prostate cancer

**DOI:** 10.1101/2025.06.06.25328738

**Authors:** Soumaya Labidi, Belinda Jiao, Shirley Tam, Parvaneh Fallah, Aida Salehi, Raghu Rajan, Mona Alameldine, Fadi Brimo, William D. Foulkes, Andreas I. Papadakis, Nabodita Kaul, Alan Spatz, Cristiano Ferrario, Ramy R. Saleh, April A. N. Rose

**Affiliations:** Gerald Bronfman Department of Oncology, McGill University, Montreal QC; Segal Cancer Center, Jewish General Hospital, Montreal QC; Lady Davis Institute for Medical Research, Jewish General Hospital, Montreal QC; Cedars Cancer Center, McGill University Health Center, Montreal QC; Department of Pathology, McGill University and Division of Pathology, Department of Clinical Laboratory Medicine, McGill University Health Center, Montreal QC; Department of Human Genetics, School of Biomedical Sciences, McGill University, Montreal QC

## Abstract

**Background:** Metastatic prostate cancers frequently harbor pathogenic aberrations in DNA damage repair (DDR) genes, that confer sensitivity to PARP inhibitors (PARPi). Therefore, accurate identification of all eligible patients is needed. The development of circulating tumor DNA (ctDNA) testing alternative is promising as genomic testing of archived tissue leads to up to 30-40% failure rate in prostate cancer.

**Methods:** This was a bi-institutional retrospective cohort study of patients with metastatic prostate cancer treated at the Jewish General Hospital or the McGill University Health Center, Montreal Canada, between 2021-23. Molecular data and treatment information was abstracted from a chart review. Chi-square, Fisher’s exact test, and Mann-Whitney tests were used to assess differences between groups.

**Results:** We identified 484 metastatic prostate cancer patients. Somatic and germline testing for DDR was performed in 55.4% (n=268) and 20% (n=97) patients, respectively. Somatic testing was performed on tissue (n=192, 71.6%) or ctDNA from liquid biopsies (n=18, 6.7%) or both (n=58, 21.7%). Pathogenic somatic DDR alterations were detected in 48 patients (17.9%). BRCA2 was the most frequent (n=17) followed by ATM (n=11), then CHEK2 (n=5). Amongst patients with germline testing 13/97 (13.4%) had pathogenic alterations predicting to lead to deficient DDR, mostly BRCA2 (n=9) and 3 had detectable BRCA2 in tissue. Dual testing modality (tissue+ctDNA) significantly enhanced the detection rate of DDR alterations 19/58 (32.7%) vs 29/210 (13.8%) for single testing modality (tissue or ctDNA) P=0.008. The rate of inconclusive results was significantly lower in dual testing modality 0/58 (0%) vs 25/210 in single testing modality (11.9%), P=0.003. Amongst the 14 patients who had discordant results between liquid and tissue tests, DDR abnormalities were more frequently identified in ctDNA (n=11) vs. tissue (n=3). Patients who had DDR deficiency detected only in ctDNA, had older tissue samples (median 5.6 years) compared to those who had deficient DDR detected only in tissue (median 0.2 years; P=0.14).

**Conclusion:** These data highlight a potential role in implementing liquid biopsy - especially in patients who only have older archival tissue available or failed tissue testing - to improve the detection rate of deficient DDR. Our ongoing prospective study will further validate whether the addition of liquid biopsy can identify more patients who are eligible to receive precision therapies.by increasing the rate of detection of DDR deficiency compared to routine tissue testing alone.

## 1. Introduction

Prostate cancer is the most commonly diagnosed cancer in males, accounting for about 1 in 5 new cancer cases (1). Despite the high long-term survival in localized prostate cancer, metastatic prostate cancer remains incurable and is the third leading cause of cancer-related mortality in men (1). Metastatic prostate cancer (mPC) is a heterogenous disease, and predictive biomarkers are increasingly being used to guide treatment decisions.

Metastatic prostate cancer can either be classified as castration-sensitive (mCSPC) or castration-resistant (mCRPC). In mCSPC, androgen deprivation therapy (ADT) is the standard primary treatment, combined with either androgen receptors pathway inhibitors (ARPI) and/or chemotherapy (2–8). Despite optimal treatment, the majority of mCSPC will eventually evolve to mCRPC, for which there is no curative treatment (9).

Deleterious aberrations or alterations in genes involved in DNA damage repair (DDR) occur frequently in metastatic prostate cancer (10–12). Pathogenic BRCA1, BRCA2 and ATM gene variants are the best characterized, and are associated with aggressive disease (10, 13–16). The PARP enzyme is essential for repairing single-strand DNA breaks. PARP inhibition results in double-strand DNA breaks which can be repaired via homologous recombination repair (HRR) mechanisms. PARP inhibition in cells with loss of function of genes that regulate homologous recombination repair thus results in excessive DNA damage that cannot be repaired and ultimately causes cell death. As such, DDR gene alterations represent important predictive biomarkers for PARP inhibitors (PARPi) such as olaparib and talazoparib in mCRPC (17, 18).

The PROfound phase 3 clinical trial randomized patients with DDR gene alterations to olaparib or physician’s choice in the second line treatment of mCRPC (19). Olaparib improved radiographic progression free survival (rPFS) and overall survival (OS) (19). The Talapro-2 trial evaluated the PARPi talazoparib plus enzalutamide vs. enzalutamide alone for the 1^st^ line treatment for mCRPC. The combination of talazoparib and enzalutamide was associated with improved rPFS and OS in an unselected population of mCRPC patients (20–22). Combinations of different PARPi with ARPI were also tested in 1^st^ line setting for mCRPC, with statistically significant benefit in rPFS (23, 24). There are ongoing trials testing different PARPi combinations in the mCSPC setting, which if positive, could change the standard of care treatment for this population of patients (25). Beyond PARPi, there is emerging data that some DDR alterations (BRCA2, ATM) may also predict benefit from carboplatin chemotherapy (26).

In Quebec, provincial reimbursement for PARPi for mCRPC is decided based on evidence of predictive biomarkers (BRCA1/2 alterations). The incidence of DDR alterations in prostate cancer data is lacking in Quebec, and little is known about testing practices and challenges in a single-payer health system. In this study, we analyzed data from two academic cancer centers in Montreal, Quebec.

The most recent guidelines recommend to perform both germline and somatic testing for all metastatic prostate cancer patients, to guide the use of PARPi, and also for germline to assess other cancer risks and counsel patients’ families (27–29). The data supporting the optimal test type to use (primary tissue, metastasis biopsy tissue or liquid biopsy) are lacking. However, the ASCO guidelines recommend considering repeat testing either on metastasis tissue biopsy or liquid biopsy, for patients with an initially negative test (27, 29).

The liquid biopsy concept was introduced for the detection of circulating tumor cells (CTC) over 10 years ago and then extended to circulating tumor DNA (ctDNA) (30, 31). CTC and ctDNA are considered as new biomarkers and subjects of translational research. Clinical applications include early cancer detection, improved cancer staging, early detection of relapse, real-time monitoring of therapeutic efficacy, and detection of therapeutic targets and resistance mechanisms (32).

A key challenge in the management of prostate cancer is accurately identifying all those patients who are eligible for new precision therapies (33). The development of ctDNA testing alternative is promising as genomic testing of archived tissue leads to up to 30-40% failure rate in prostate cancer. This can be due to low cellularity in fine needle biopsy specimens for pathological analysis, or to limited quality or quantity of DNA due to degradation over time or both (33). Repeating biopsies on metastatic sites, often bone metastases, are invasive time-consuming and resource-consuming procedures, which are often futile due to the lack or poor quality of tissue obtained, especially that the decalcification process degrades DNA and leads to NGS failures.

We sought to determine whether liquid biopsy combined with tissue testing resulted in a higher rate of detection of DDR gene alterations for patients with metastatic prostate cancer.

## 2. Patients and Methods

### 2.1. Patient population, characteristics and outcome

We performed a bi-institutional retrospective cohort study analysis of patients treated for metastatic prostate cancer at two Canadian cancer centers in Montreal, Quebec: Segal Cancer Center, Jewish General Hospital (JGH) and Cedars Cancer Center, McGill University Health Center (MUHC). Patients with proven metastatic prostate cancer, with active follow-up in medical oncology clinics between January 2021 and December 2023 were included. Clinical, pathologic and molecular characteristics were abstracted from chart review. DDR alterations were considered when they were reported as pathogenic or likely pathogenic variants.

The following characteristics were collected: age, cancer history, pathological findings, stage of disease at the time of diagnosis, site of metastases, and treatment type at localized and metastatic stage. Types of molecular testing, access, time from testing request to results and detailed results were assessed. We collected the following outcome criteria: investigator assessed clinical and radiographic response, PSA50 response, time to castration resistance, and OS. Time to castration resistance was calculated from the diagnosis of metastatic disease to castration resistance. OS was calculated from the diagnosis of metastatic disease to death or last follow-up, and from the diagnosis of castration resistance to death or last follow-up. Clinical data were stored in a RedCap database.

### 2.2. Statistical analysis

Descriptive statistics are provided for patient characteristics. Chi-square, Fisher’s exact test, and Mann-Whitney tests were used to assess differences between groups. The figures were created using the GraphPad Prism version 10.4.1.627 for Windows, GraphPad Software (www.graphpad.com).

### 2.3. Ethics approval

Ethics approval for the study was obtained from the Integrated University Health And Social Services Centres (CIUSSS) West Central Montreal REB (Project MP-05-2024-3885) on January 12, 2024.

## 3. Results

### 3.1. Patients’ characteristics

We identified 484 patients who were treated for mPC in medical oncology clinics between 2021 and 2023. The median age was 67 years (42–92). Most of the patients (n=453, 93.6%) were diagnosed with adenocarcinoma. The Gleason score was ≥ 8 in 61.6% (n=298) of the cases. More than half of the patients were diagnosed with de novo metastatic disease (n=253, 52.3%). At diagnosis of metastatic disease, 49.2% and 38,8% of the patients had high volume and high risk disease respectively. Median follow-up from the time of diagnosis was 6.6 years, and 3.8 years from the diagnosis of metastatic disease. Patients’ characteristics for the entire cohort are shown in **Table 1**.

**Table 1:** Patients’ characteristics.

|  |  |  |
| --- | --- | --- |
| N = 484 |  |  |
| **Centre** |  | N (%) |
|  | JGH | 240 (49.6) |
|  | MUHC | 244 (50.4) |
| **Age Years** | Median (Range) | 67 (42 – 92) |
| **Medical History** |  | N (%) |
| Personal history of cancer | Yes | 80 (16.5) |
|  | No | 396 (81.8) |
|  | Missing data | 8 (1.7) |
| Family history of cancer | Yes | 187 (38.6) |
|  | No | 173 (35.8) |
|  | Missing data | 124 (25.6) |
| **Stage at diagnosis** |  | N (%) |
|  | Localized | 230 (47.5) |
|  | Metastatic | 253 (52.3) |
|  | Missing data | 1 (0.2) |
| **Pathology** |  | N (%) |
| Pathology | Adenocarcinoma | 453 (93.6) |
|  | Other | 3 (0.6) |
|  | Missing Data | 28 (5.8) |
| Gleason | 7 | 112 (23.1) |
|  | 8 | 80 (16.6) |
|  | 9 | 189 (39) |
|  | 10 | 29 (6) |
|  | Missing data | 74 (15.3) |
| **Treatment for localized disease** |  | N (%) * |
|  | Surgery | 98 (20.2) |
|  | Radiation | 238 (49.2) |
|  | ADT | 104 (21.5) |
|  | Adjuvant ARPI (Abiraterone) | 5 (1) |
|  | Adjuvant chemotherapy (Docetaxel) | 4 (0.8) |
|  | Not applicable | 182 (37.6) |
|  | Clinical trial | 5 (1) |
|  | Missing data | 13 (2.7) |
| **Site of metastases at diagnosis of metastatic disease** |  | N (%) ** |
|  | Bone | 382 (79.5) |
|  | Lymph Nodes | 194 (40.4) |
|  | Visceral | 56 (11.7) |
|  | Other | 7 (1.5) |
|  | Missing data | 5 (1) |
| **Disease characteristics** |  | N (%) |
| High volume as per CHAARTED trial | Yes | 236 (49.2) |
|  | No | 211 (43.9) |
|  | Missing data | 33 (6.9) |
| High risk as per LATITUDE trial | Yes | 186 (38.8) |
|  | No | 225 (46.9) |
|  | Missing data | 69 (14.4) |
| **Castration resistance** |  | N (%) |
|  | Yes | 332 (68.6) |
|  | No | 152 (31.4) |

\*Sums do not add to 100% as one patient may have received multiple treatment modalities; \*\*Sums do not add to 100% as one patient may have multiple sites of metastases

### 3.2. Germline molecular testing

Germline testing was performed in 20% (n=97) of the cases. Most of the patients who had germline testing also had a personal and/or family history of cancer (n=83), including prostate, breast and gastro-intestinal cancers (**Supplemental Table 1**). The majority of the germline testing was done locally (n=56, 57.7%) through consultation with Medical Genetics. Details are shown in **Table 2**. The median time for results from the date of testing request by the clinician was 1.6 months (0.2 – 9.7). Pathogenic DDR alterations were identified in 13 patients (13.4%). BRCA2 was the most frequent (n= 9, 69.2 %), followed by CHEK2 (n= 2, 15.4%). One patient had 2 alterations in BRCA2 and PALB2 genes (**Table 2**).

**Table 2:** Germline molecular testing.

| N = 484 |  | N (%) |
| --- | --- | --- |
| Germline testing done | Yes | 97 (20) |
|  | No | 386 (79.8) |
|  | Missing data | 1 (0.2) |
| Testing access | Local | 56 (57.7) |
|  | Trial | 20 (20.6) |
|  | Access program | 15 (15.5) |
|  | Other | 4 (4.1) |
|  | Missing data | 2 (2.1) |
| Pathogenic DDR Present | Yes | 13 (13.4) |
|  | No | 83 (85.6) |
|  | Missing data | 1 (1) |
| DDR Alterations | BRCA2 | 9 (69.2) |
|  | CHEK2 | 2 (15.4) |
|  | RAD51C | 1 (7.7) |
|  | BRCA2+PALB2 | 1 (7.7) |

### 3.3. Somatic molecular testing

Somatic testing was performed for 268/484 (55.4%) patients (**Table 3**). Somatic testing was principally performed locally at the JGH pathology department (n=193, 72%). The testing was performed by a single modality, either tissue or ctDNA in 71.6% (n=192) and 6.7% (n=18), respectively. Dual modality testing of both tumor tissue and ctDNA was performed for 58 patients (21.7%). Detailed results are shown in **Table 3**. The median time from testing request by the clinician to the testing results was shorter for ctDNA testing 0.73 months (0.33 – 4.43) vs 1.36 months (0.06 – 13.2) for tissue testing (p=0.0006) (**Table 4**).

**Table 3:** Somatic molecular testing access and type.

|  |  |  |
| --- | --- | --- |
| N = 484 |  | N (%) |
| Somatic testing done | Yes | 268 (55.4) |
|  | No | 216 (44.6) |
| Testing access* | Local | 193 (72) |
|  | Trial | 68 (25.4) |
|  | Access program | 35 (13) |
|  | Other | 5 (1.8) |
| Testing type | Tissue | 192 (71.6) |
|  | ctDNA | 18 (6.7) |
|  | Both | 58 (21.7) |

\*Sum does not equal 100% as some patients can have multiple tests

**Table 4:** Somatic molecular testing: Tissue vs ctDNA.

|  |  | Tissue N (%) | ctDNA N (%) | P-Value |
| --- | --- | --- | --- | --- |
| Conclusive | Yes | 215 (86) | 68 (88.3) | 0.48 |
|  | No | 31 (12.4) | 7 (9.1) |  |
|  | Missing data | 4 (1.6) | 2 (2.6) |  |
| Time to testing results (months) | Median (Range) | 1.36 (0.06 – 13.2) | 0.73 (0.33 – 4.43) | 0.0006 |

Somatic testing was reported as conclusive in most cases, regardless of the testing modality: 86% for tissue and 88.3% for ctDNA (**Table 4**). Amongst patients who had somatic testing performed, DDR alterations were identified in 48 (17.9%). BRCA2 alterations occurred most frequently (n=17, 35.4%), followed by ATM (n=11, 22.9%) then CHEK2 (n=5, 10.4%) (**Figure 1**). Among the 13 patients with identified germline pathogenic DDR alteration, 7 underwent somatic testing. The pathogenic alteration was concordant in both testing modalities in 6/7 (85%) cases: BRCA2 (n=3), CDK12 (n=1), RAD51C (n=1) and BRCA2+PALB2 (n=1).

**Figure 1:**
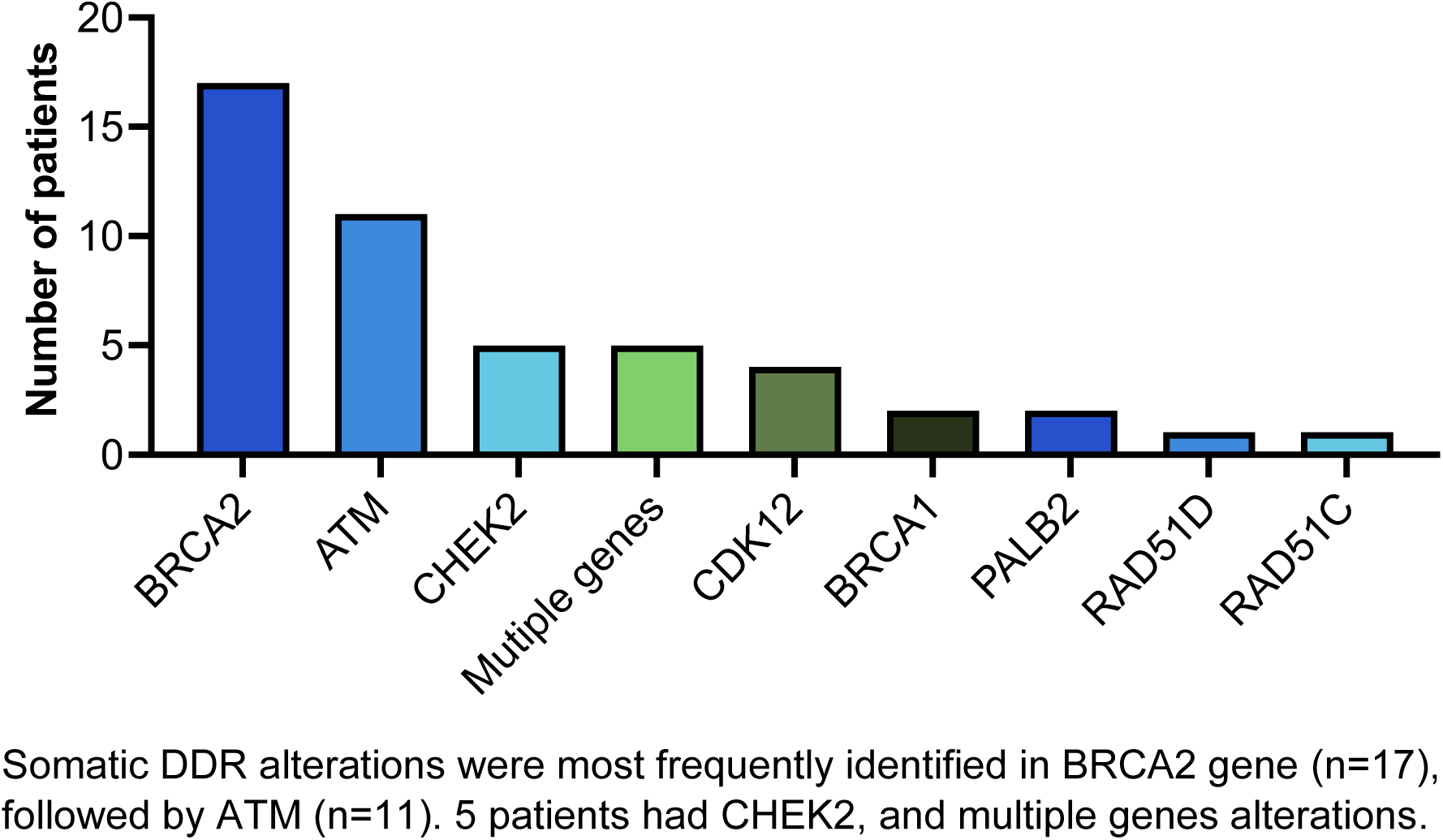
Somatic DDR Alterations. Somatic DDR alterations were most frequently identified in BRCA2 gene (n=17), followed by ATM (n=11). 5 patients had CHEK2, and multiple genes alterations.

### 3.4. Single vs dual modality somatic testing

In this cohort, there were 58 patients who had both tissue and ctDNA analysis. The DDR alteration detection rate was significantly enhanced in these patients (19/58; 32.7%) compared to patients who had only one test – either tissue or ctDNA (29/210; 13.8%) p=0.008; or compared to patients who had only tissue testing (27/192; 14%; p=0.010) (**Table 5**). The rate of inconclusive results was significantly lower for patients with dual modality testing 0/58 (0%) vs single modality testing 25/210 (11.9%), p=0.003 (**Table 5**). Poor DNA quality was the most common reason for an inconclusive testing result. (**Table 6**).

**Table 5:**
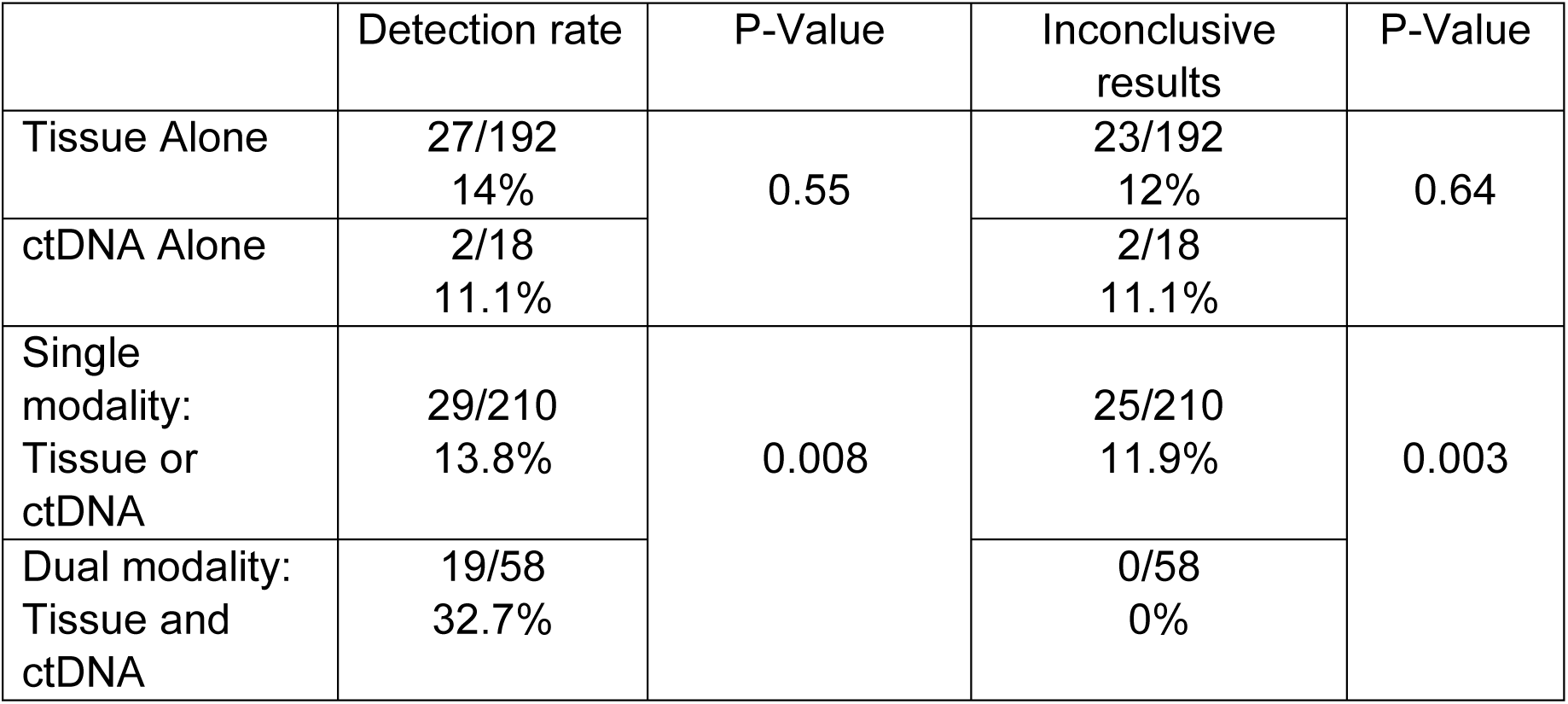
Detection rate and inconclusive results according to testing modality.

**Table 6:** Inconclusive results on tissue testing.

| Reason | N (%) | Details |
| --- | --- | --- |
| Poor DNA quality | 17 (73.9) | Median age of tissue at the time of analysis<br>Years (Range) 8 (2-20) |
| Tissue not available | 6 (26.1) |  |

Among the patients who had dual modality testing with both tissue and ctDNA analysis, we observed discordant results in 14 cases (**Table 7**). Three patients with tissue positive and negative ctDNA, were tested at diagnosis of mCSPC, with blood collected 2 to 4 weeks after starting ADT treatment for 2 of them. Conversely, for 11 patients, tissue results were negative and ctDNA positive. The median age of tissue at the time of testing was 5.6 years compared to 0.2 years in the tissue positive ctDNA negative cases.

**Table 7:** Discordant results between tissue and ctDNA.

| Patient | Stage at the time of analysis | Tissue conclusive | ctDNA conclusive | DDR | Tissue results | ctDNA results | Tissue age at time of analysis (Years) | Median Age (Years) | P-Value |
| --- | --- | --- | --- | --- | --- | --- | --- | --- | --- |
| JGH026 | mCSPC | Yes | Yes | CDK12 | + | - | 0.2 | 0.2<br>(0.1 – 4) | 0.1236 |
| JGH028 | mCSPC | Yes | Yes | BRCA1 | + | - | 0.1 |  |  |
| JGH031 | mCSPC | Yes | Yes | PALB2 | + | - | 4.0 |  |  |
| JGH022 | mCRPC | No* | Yes | CHEK2 | - | + | 10.0 | 5.6<br>(0.2 – 12.3) |  |
| JGH029 | mCRPC | Yes | Yes | CDK12 | - | + | 2.5 |  |  |
| JGH030 | mCSPC | Yes | Yes | ATM | - | + | 7.0 |  |  |
| JGH032 | mCRPC | Yes | Yes | BRCA2 | - | + | 2.5 |  |  |
| JGH033 | mCRPC | Yes | Yes | BRCA2<br>PALB2 | - | + | 6.0 |  |  |
| JGH034 | mCRPC | Yes | Yes | CHEK2 | - | + | 12.3 |  |  |
| JGH035 | mCRPC | No* | Yes | BRCA2 | - | + | 11.0 |  |  |
| JGH036 | mCRPC | Yes | Yes | ATM | - | + | 3.3 |  |  |
| JGH164 | mCRPC | Yes | Yes | ATM | - | + | 5.6 |  |  |
| JGH168 | mCRPC | Yes | Yes | CHEK2<br>CDK12 | - | + | 0.2 |  |  |
| RVH133 | mCSPC | Yes | Yes | ATM | - | + | 0.2 |  |  |

\*Poor DNA quality

### 3.5. Treatment with PARPi: Results and outcomes

Within the entire cohort, 28 patients received a treatment with PARPi. The treatment was received as standard of care in 57.1% of the cases (n=16), mostly in the mCRPC setting (n=26, 92.9%). Detailed results are shown in **Table 8**. A PSA50 response in mCRPC was obtained in 34.6% of the cases (n=9).

**Table 8:** Treatment with PARP inhibitors.

| N = 28 |  | N (%) |
| --- | --- | --- |
| DDR Alteration | Germline BRCA2 | 2 |
|  | Somatic BRCA2 | 9 |
|  | Somatic BRCA1 | 2 |
|  | Somatic ATM | 2 |
|  | Somatic CHEK2 | 2 |
|  | Somatic CDK12 | 1 |
|  | Somatic PALB2 | 1 |
|  | Somatic BRCA2+PALB2 | 1 |
|  | Somatic and Germline BRCA2 | 2 |
|  | Somatic and Germline BRCA2+CDK12 | 1 |
| Type of treatment | Blinded randomized trial | 7 (25) |
|  | Open label trial | 5 (17.9) |
|  | Standard of care | 16 (57.1) |
| Stage | mCSPC | 2 (7.1) |
|  | mCRPC | 26 (92.9) |
| Line of treatment for mCRPC | First | 9 (34.6) |
|  | Second | 5 (19.2) |
|  | Third | 8 (30.8) |
|  | ≥ Fourth | 4 (15.4) |
| PSA50 response in mCRPC | Yes | 9 (34.6) |
|  | No | 13 (50) |
|  | Missing data | 4 (15.4) |

Three patients with DDR alteration detected on ctDNA with negative tissue results, were able to receive PARPi treatment as per standard of care, with response achieved in 2/3 cases (**Table 9**).

**Table 9:** Access to PARPi for patients with discordant results between tissue and ctDNA.

| Patient | DDR | Tissue results | ctDNA results | PARPi Treatment | Stage | Treatment Line | Duration of Treatment | PSA50 Response |
| --- | --- | --- | --- | --- | --- | --- | --- | --- |
| JGH032 | BRCA2 | - | + | olaparib | mCRPC | 4<sup>th</sup> Line | 3 Months | No |
| JGH033 | BRCA2<br>PALB2 | - | + | olaparib | mCRPC | 3<sup>rd</sup> Line | 1.8 years;<br>on-going | Yes |
| JGH035 | BRCA2 | - | + | olaparib | mCRPC | 2<sup>nd</sup> Line* | 3.9 years;<br>on-going | Yes |

\*Small cell differentiation at progression. PARPi post Etoposide-Platinum chemotherapy

## 4. Discussion

Pathogenic DDR alterations occur frequently in prostate cancer. In one study, biallelic inactivation of BRCA2, BRCA1 or ATM was observed in nearly 20% of the affected individuals (11). In a multicentre study on 692 men with mPC, the incidence of pathogenic germline alterations in DDR pathway was 11.8% (12). Mutation frequencies did not differ according to whether a family history of prostate cancer was present or according to age at diagnosis (12). In our bi-institutional retrospective cohort study analysis of patients treated for mPC, we report an incidence of 13.4% for germline pathogenic DDR alterations, and 17.9% for somatic DDR alterations. BRCA2 was the most frequent in both germline and somatic, 69.2% and 35.4% respectively.

Genomic alterations in DDR pathway, and especially BRCA1/2 deficient mPC, show a more aggressive phenotype and poor survival outcomes (13–16, 34). Apart from the prognostic value, somatic and germline alterations also have a predictive value of response to PARPi and platinum based chemotherapy, which affects treatment decision making for patients with mPC (17–20, 23, 24, 26, 34–36). Germline genetic testing may also offer hereditary cancer risk information requiring genetic counseling (12, 27, 29). Therefore, the ASCO guidelines, (27, 29), and the Canadian Urological Association guidelines (28) recommend germline and somatic testing for all mPC. Little is known about the current testing practices in the context of the Canadian healthcare system. In 2022, a cross-sectional survey was led by the Canadian Genitourinary Research Consortium (GURC), to an academic multi-disciplinary group of investigators across 22 GURC sites (37). The results showed that 84% of the investigators were offering genomic testing to patients with advanced prostate cancer, mostly germline (94% compared to 72% on tissue) (37). Here, we report real-world data for testing practices in two academic centers in Montreal, Quebec. Despite the availability of local tissue testing and genetic consultation, only 55.4% and 20% of the patients were offered somatic and germline testing, respectively. Importantly, provincial reimbursement for olaparib was first approved in Quebec in 2022, which likely influenced local practice patterns.

Numerous guidelines recommend genetic testing in prostate cancer, however there is a lack of consensus regarding who to test and how the tests should be performed (38, 39). The recommendations used to range from targeted gene test for 1 or 2 genes to a prespecified gene panel (38). The most recently published ASCO guidelines recommend next-generation DNA sequencing panel-based assays (NGS) (27, 29). The NGS-based tests include multiple genes associated with cancer risk factors (40). The NGS panels are customizable and allow the selection of actionable genes for specific testing purposes such as the DDR pathway genes (39, 40). Tissue is a reliable option and remains the gold standard sample for somatic testing in prostate cancer (39, 40). However, it has some limitations and faces challenges. In fact, the isolation of an evaluable quality and quantity of DNA depends on multiple pre-analytical and analytical factors, such as biopsy route, biopsy technique, tissue processing and storage of Formalin-Fixed Paraffin-Embedded (FFPE) blocks (39, 41). The tumor heterogeneity might result in missing late somatic mutations especially if testing is conducted on archival sample (39, 42). There is a higher prevalence of DDR gene alterations in metastatic vs. primary tumor samples (19), but obtaining tissue samples from metastatic sites can be challenging in mPC, as the most frequent site of metastasis is bone and isolation of DNA from bone require specific decalcification protocols that may degrade the quality of the DNA (39, 43). In the PROFOUND clinical trial, the rate of tissue test failure was 31% from predominantly archival FFPE tissue samples, with DNA extraction failure as the most frequent cause (13.2%) (19, 44). In our cohort, the rate of inconclusive results on tissue was 12%, with poor DNA quality as the most frequent reason for failure (73.9%). The median age of these samples at the time of analysis was 8 years (2–20). It is proven that the quality of the FFPE samples and the storage conditions can affect the quality of the DNA (44–46). Liquid biopsy and testing on ctDNA offers a valid minimally invasive alternative to tissue testing. Evaluating ctDNA can also provide an overall view of tumor heterogeneity and emerging genomic alterations (32, 47, 48). Serial ctDNA testing in mCRPC identify 11% of new actionable alterations, with 30% of all BRCA2 alterations identified only on repeat testing (48). The limitations from liquid biopsy include false negatives from low tumor burden and variations according to treatment phase (39, 49). Clonal hematopoiesis of indeterminate potential (CHIP) variants detected on both plasma and whole blood are a known confounder of ctDNA testing (50–52). Paired whole-blood control testing allows to distinguish between CHIP and prostate cancer variants (51). The ctDNA testing in our study was performed mostly through access programs and clinical trials, with different techniques, therefore we could not properly assess whether some of the DDR alterations identified in cfDNA were derived from CHIP rather than from prostate cancer ctDNA.

We observed a higher DDR gene alteration detection rate in patients who had dual modality testing compared to single modality, and significantly lower inconclusive results. Chi et al. reported an 81% positive percentage agreement and 92% negative percentage agreement for BRCA and ATM status on tissue compared with matched ctDNA samples from the Profound trial cohort (53). Discordant results with positive BRCA or ATM alteration on tissue and negative ctDNA were found in 19% of the cases (53). Non evaluable ctDNA fraction or low ctDNA fractions when evaluable were enriched in these cases, possibly related to low tumor burden (53). The three cases with discordant results in our cohort were tested in the mCSPC setting shortly after treatment starts, which could lead to low ctDNA fraction. Tissue and ctDNA testing both have limitations and cannot capture all the actionable alterations in all patients, but the availability of both enhances the detection rate of relevant alterations and offers our patients efficient treatment options. We report 2 cases with negative tissue testing and positive ctDNA serial testing for BRCA2 and BRCA2+PALB2 alterations, who achieved sustained response on PARPi. This illustrates the importance of accurate identification of patients eligible to precision therapies.

## 5. Conclusion

DDR pathway alterations are major prognostic and predictive factors in mPC. Somatic and germline NGS testing is recommended for all mPC to guide treatment planning, but also to guide eventual genetic counseling and cascade testing (27, 29). Tissue testing remains the preferred option; however, it can be challenging in prostate cancer due to limitations from old archival samples, re-biopsy possibilities especially on bone metastases. ctDNA is a valid alternative, and our data concludes to an enhanced detection rate and significantly lower inconclusive results with combining tissue and ctDNA testing. Our study has some limitations, especially the small size and the retrospective character. However, it is hypothesis generating, therefore we are currently conducting a prospective research project, where all mPC patients will have dual testing modalities, to validate this hypothesis.

## Data Availability

All data produced in the present work are contained in the manuscript

